# Evaluation of gastric electrophysiology, symptoms and quality of life after pancreaticoduodenectomy

**DOI:** 10.1101/2023.12.14.23299984

**Authors:** Tim Hsu-Han Wang, Chris Varghese, Stefan Calder, Armen Gharibans, Gabriel Schamberg, Adam Bartlett, Sanket Srinivasa, Greg O’Grady

## Abstract

**Background:** Pancreaticoduodenectomy (PD) is an operation performed for several indications, including pancreatic and biliary malignancies. Delayed gastric emptying (DGE) is a common post-operative complication and the underlying pathophysiology remains poorly understood. This study aimed to evaluate the gastric electrophysiology, symptoms and quality of life following PD, using the novel non-invasive Gastric Alimetry System.

**Methods:** PD patients with index operations between 2017-2022 were assessed using the Gastric Alimetry System®, comprising a stretchable 8×8 electrode array, wearable Reader, and validated symptom logging app. A 30-minute pre-prandial and a 4-hour post-prandial recording was performed. Outcomes included Principal Gastric Frequency, BMI-adjusted amplitude, Gastric Alimetry Rhythm Index, meal response, symptoms, and quality of life (QOL) questionnaires (PAGI-SYM, PAGI-QOL). Data was compared to a validated reference range and matched healthy controls.

**Results:** 19 patients and 19 matched controls were recruited. There were no differences in any gastric parameters between PD patients and matched controls (all *p*>0.05). Gastric electrophysiology parameters lay outside normative reference intervals in 8/19 cases, generally being only minor deviations, while significant symptoms occurred in 8/19 (42%); which did not correlate (*p*=0.43). PD patients had worse quality of life scores than controls (p<0.01), however, no correlations were identified between electrophysiological parameters and QOL.

**Discussion:** Moderate to severe upper GI symptoms are common after PD with worse QOL compared to the controls. Gastric electrical activity generally recovers well long-term following PD, indicating that other factors contribute to symptoms. Further studies should now assess acute changes in gastric function after PD.

## Introduction

Pancreaticoduodenectomy (PD) is a complex hepatopancreatobiliary (HPB) procedure performed for both malignant and benign conditions, with the most common indication for pancreatic or bile duct malignancies. Operatively, these procedures typically involve the resection of distal aspect the stomach, and anastomosing the distal stomach onto a selected section of the small intestine with reconstruction (eg. Roux-en-Y, Billroth II reconstruction). Post-operative complications are common, including pancreatic fistula and delayed gastric emptying (DGE), which ranges between 10 to 45%, along with non-specific symptoms of nausea, vomiting and abdominal pain, which impairs quality of life (QoL) and prolongs hospitalisation, as well as increasing healthcare costs (1–4). While there have been several operative attempts at reducing the incidence of DGE (antecolic vs retrocolic, pylorus-preserving vs pylorus-resecting), the underlying pathophysiology is still to be determined.

Advances in the understanding of gastric electrophysiology and motility has led to gastric dysrhythmias being implicated in conditions such as gastroparesis and chronic unexplained nausea and vomiting, as well as chronic post-operative symptoms (5, 6). More recently, an aberrant gastric conduction pathway at the site of small bowel anastomosis has been found in post-surgical gastric dysfunction (7).

Gastric Alimetry® (Auckland, New Zealand) is a new non-invasive test to evaluate gastric electrophysiology and function at high resolution (HR), recently receiving regulatory approvals for clinical use (8). This technique has been extensively validated (5, 7, 9), and is being applied in medical disorders and more recently on esophagectomies, but has yet to be used for post-PD patients (10). This study aimed to assess the electrophysiology of the post-PD stomach using the Gastric Alimetry system, together with symptom evaluation and quality of life in post-PD patients.

## Methods

### Patient population

Patients who underwent PD between 2017 and 2022 in the Auckland region (New Zealand) were recruited. Ethical approval was obtained from the Auckland Health Research Ethics Committee (AH1125) and informed consent was obtained from all patients. Patients who had a history of skin allergy, evidence of mechanical gastric or small bowel obstruction as a cause for their symptoms, those without a post-operative CT scan, and those who have insulin-dependent diabetes were excluded. Clinical data including operation notes, imaging, endoscopy, and histopathology (if applicable) were evaluated. Patients were also followed up within 1 week for adverse reactions. A healthy control cohort was also recruited and matched based on sex and BMI.

### Quality of Life Assessments

At the time of the Gastric Alimetry, validated quality of life assessment questionnaires were used including the PAGI-SYM, PAGI-QOL and EQ-5D-5L, which assess symptoms and quality of life over the preceding 2 weeks. Individual and total scores were obtained and analysed.

### Gastric Alimetry Test Methodology

Gastric Alimetry was performed under a protocol adapted for patients with a reduced gastric volume, with customised array placement based on the location of the stomach on the most recent cross sectional imaging (10). This device comprises an HR stretchable electrode array (8×8 electrodes; 20 mm spacing; 196 cm^2^), a wearable Reader, validated iOS app for symptom logging, and a cloud-based reporting platform (**Figure 1**) (11–13). Baseline recordings were performed in the first 30 minutes, followed by a 218kCal meal, comprising of 100mL of Ensure (93kCal; Abbott Nutrition, IL, USA) and half an oatmeal energy bar (125kcal, 2.5g fat, 22.5g carbohydrate, 5g protein, 3.5g fibre; Clif Bar & Company, CA, USA) to account for the partial loss of stomach reservoir, consumed over 10 minutes and a 4-hr postprandial recording in order to capture a full gastric activity cycle. Patients were seated in a chair and asked to limit movement, talking, and sleeping, but were able to read, watch media, work on a mobile device, and mobilize for comfort breaks. Symptom development or changes were recorded on the Gastric Alimetry App, calculated into a Total Symptom Burden Score (TSBS).

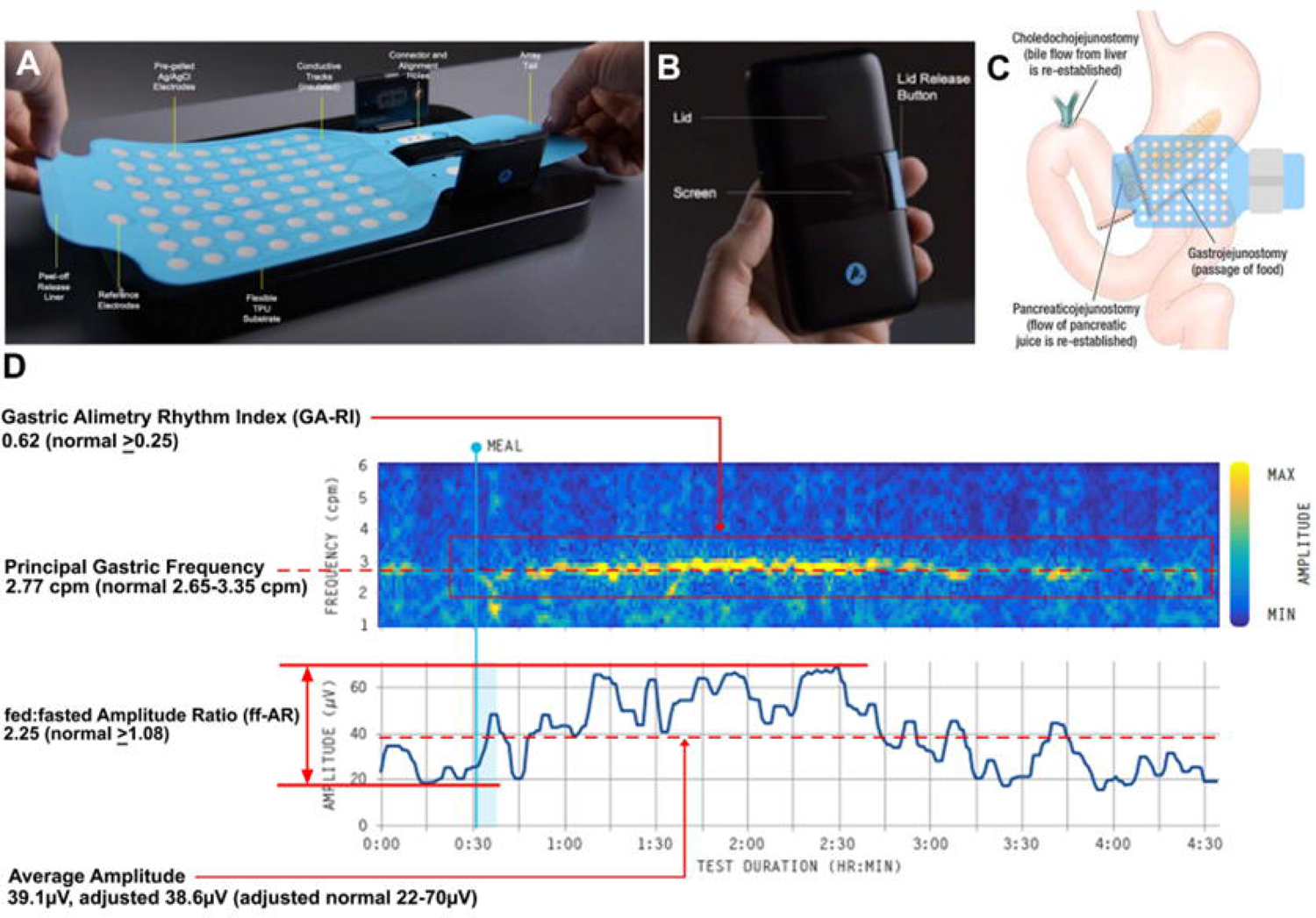

### Gastric Alimetry Data Analysis

Gastric Alimetry data analysis comprises two components: spectral analysis and symptom phenotyping (14). Spectral analysis encompasses four established metrics (15): Principal Gastric Frequency (the dominant frequency at which gastric activity is recorded), BMI-Adjusted Amplitude, Gastric Alimetry Rhythm Index (GA-RI; reflecting pacemaker stability), fed:fasted Amplitude Ratio (ff-AR; indicating meal response with contractions) (**Figure 1**). These metrics were assessed with comparison to established reference intervals from a sample of 110 healthy controls (16), as well as the matched control cohort for group-level analytics. Frequency was not reported if there was no identifiable rhythm (as measured by GA-RI) (11).

Gastric Alimetry symptom phenotyping was performed according to an emerging scheme developed by the Body Surface Gastric Mapping Working Group (14). The following categories were recognised, which can aid in the identification of symptom origins within the GI tract:

- *Sensorimotor*: symptom profile in which post-prandial symptom curves are highly correlated with gastric activity (r=0.5), indicating an association between gastric activity and symptoms (17, 18).
- *Continuous*: symptom profile that does not correlate with either the meal or the gastric activity curve. This symptom profile has been more commonly associated with gut-brain axis disorders and neuropathy (8, 19).
- *Post-gastric*: symptom profile that increases after the gastric activity curve has diminished, indicating that the symptoms arise after the gastric meal response, therefore more likely to be of small-bowel origin (14, 17).

### Safety

Patients were followed up in 1 day and 1 week and all adverse events arising from Gastric Alimetry device use were recorded.

### Data Analyses

Statistical analysis was performed using GraphPad Prism (San Diego, CA, USA) and R v.4.0.1 (R Foundation for Statistical Computing, Vienna, Austria). Correlation analysis between spectral metrics, TSBS and QoL scores were performed with Spearman’s correlation with p<0.05 as statistically significant. Data was generated on a correlation matrix then presented in a wheel plot. Spectral analysis comparisons with the age and gender matched healthy control patients were performed using the unpaired t-test with p<0.05 designated as statistically significant.

## Results

19 patients were recruited (13 males and 6 females; median age 65 years; range 35-85), together with an equal number of matched controls. PD were performed with a median of 36 months (range 10-62 months) prior with a combination of gastrojejunostomy (n=16) and Roux-en-Y (n=3) reconstructions. 18 patients had a peri-antral transection and with 1 patient having a transection distal to the pylorus. Patients and matched control demographics are presented in **Table 1**; there were no significant difference between the BMI or sex, although a modest difference was noted with regard to age (p=0.02).

| Characteristics | Patients (n=19) | Matched controls (n=19) | p value |
| --- | --- | --- | --- |
| Age | 63.8 ± 4.6 | 55.1 ± 8.6 | 0.02 |
| BMI (kg/m<sup>2</sup>) | 26.2 ± 12.4 | 55.1 ± 10.5 | 0.95 |
| Gender |  |  |  |
| Male | 13 | 9 | 0.32 |
| Female | 6 | 10 |  |
| Duration since surgery (months) | 34.9 ± 18.2 | N/A |  |
| **QOL** |  |  |  |
| TSBS | 6.3 ± 7.0 | 0.8 ± 2.4 | <0.01 |
| PAGI-SYM | 17 ± 16 | 9 ± 17 | 0.15 |
| PAGI-QOL | 30 ± 29 | 136 ± 22 | <0.001 |
| EQ-5D-5L | 0.89 ± 0.14 | 0.97 ± 0.08 | 0.07 |
| Self-reported EQ-5D-5L | 77 ± 11 | 89 ± 7 | <0.001 |
| **Spectral Characteristics** |  |  |  |
| Frequency (2.65 - 3.35cpm) | 3.03 ± 0.23 | 3.09 ± 0.19 | 0.36 |
| Adjusted amplitude (22-70μV) | 45.4 ± 20.0 | 38.6 ± 16.9 | 0.27 |
| ff-AR (≥1.08) | 1.68 ± 0.62 | 2.02 ± 0.99 | 0.24 |
| GA-RI (≥0.25) | 0.49 ± 0.22 | 0.52 ± 0.21 | 0.73 |

### Quality of Life Assessments

QoL data and TSBS are also presented in **Table 1**. Compared to the control group, the PD patients reported worse TSBS (6.3 ± 7.0 vs 0.8 ± 2.4; p<0.01) and substantially lower PAGI-QOL scores (30 ± 29 vs 136 ± 22; p<0.001) and a lower self-reported EQ-5D-5L score (77 ± 11 vs 89 ±7; p<0.001; rated on a 100 point sliding scale). There were 8/19 patients who had TSBS≥5, of which 5/8 had TSBS≥10.

### Gastric Alimetry

#### Spectral Analysis

Gastric activity was successfully captured in all cases. 8/19 cases had at least one gastric slow wave spectral parameter that lay outside the normative reference range, including one patient who had abnormal metrics in both GA-RI and Principal Gastric Frequency (**Figure 2A**, **Figure 3B**). However, most of these abnormalities lay just beyond normative reference intervals, indicating most of these physiological deviations were mild (**Figure 2D**). With regard to frequency, 2/19 cases showed mildly elevated Principal Gastric Frequency compared to the reference range data (e.g. **Figure 2B**). There were also 3 cases which were found to have mildly raised BMI-adjusted amplitude, while only one demonstrated a borderline low amplitude. When compared to the matched controls, no statistical differences were found for all spectral characteristics (p>0.20).

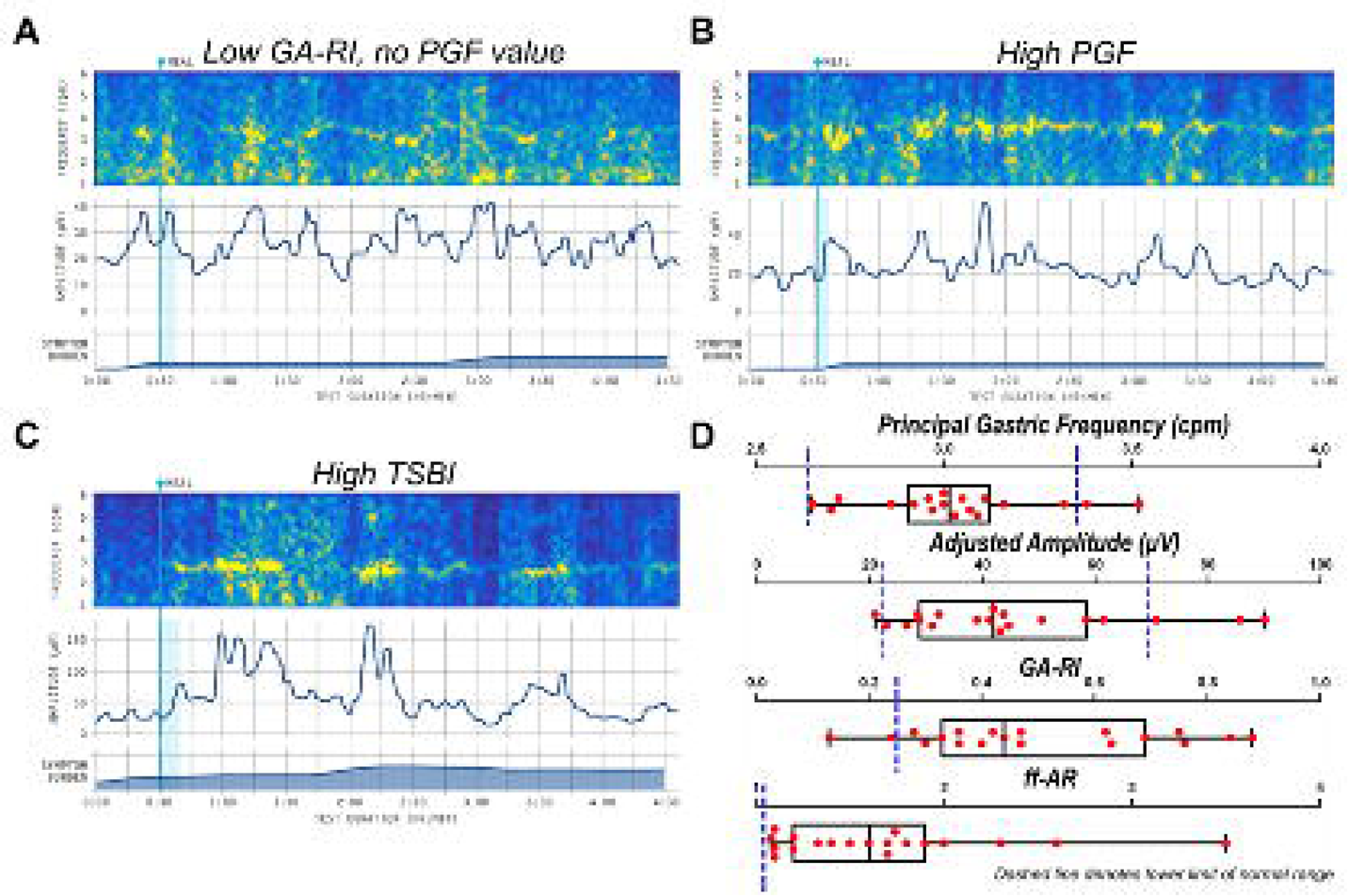

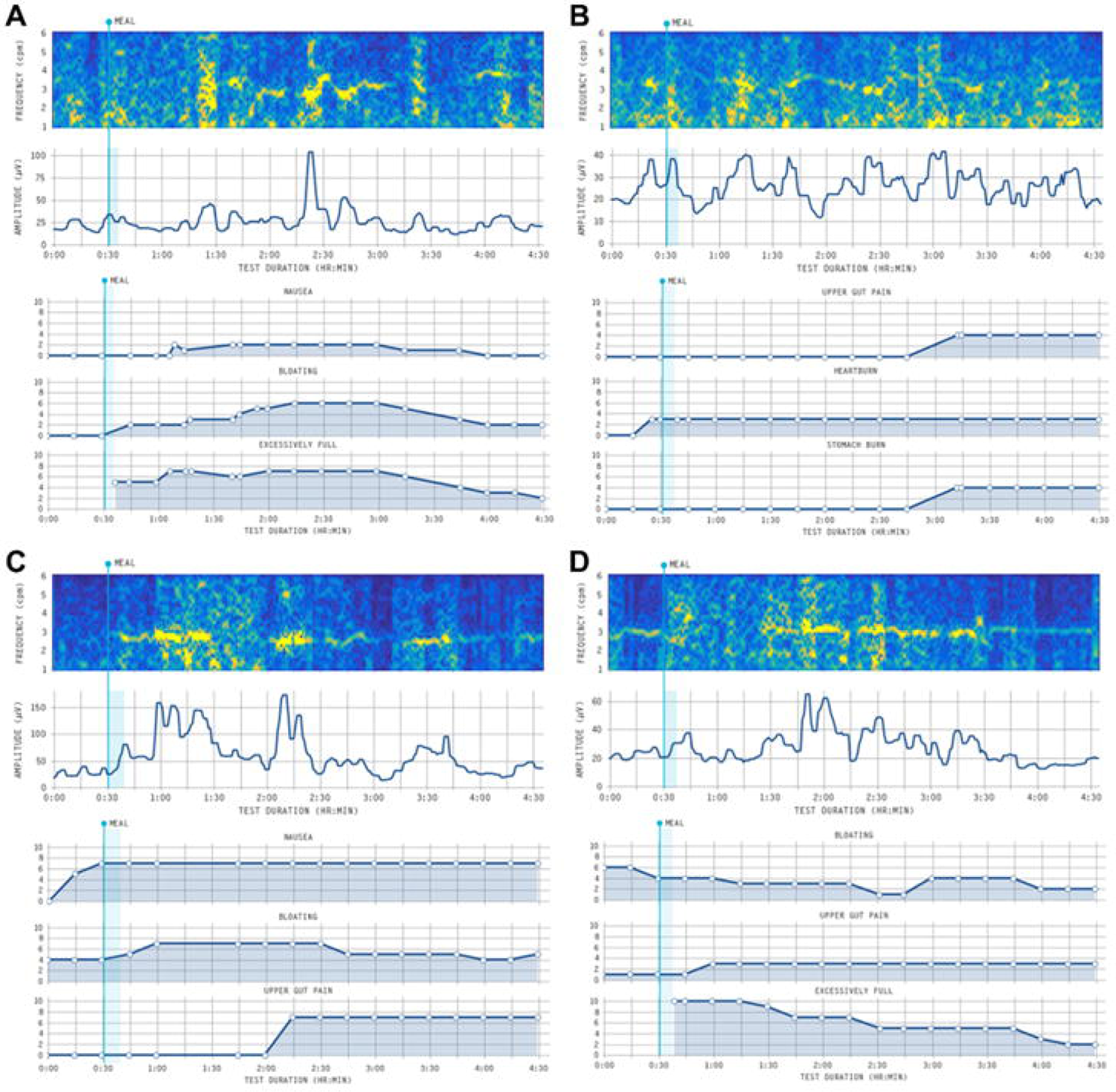

#### Symptom phenotype analysis

Gastric Alimetry symptom phenotypes observed in the post-PD patients were variable. Of the 7 patients with significant symptom burdens occurring chronically after PD, these symptom patterns encompassed a combination of sensorimotor (n=1), post-gastric (n=1), continuous (n=2) and mixed (n=3) phenotypes with examples presented in **Figure 3**. As demonstrated on the wheel-plot in Figure 4, there were minimal correlations observed between spectral metrics and patient QoL or TSBS. 3 patients who had TSBS>5 concurrently had abnormal spectral characteristics, including abnormal GA-RI and PGF, and abnormal BMI-adjusted amplitude.

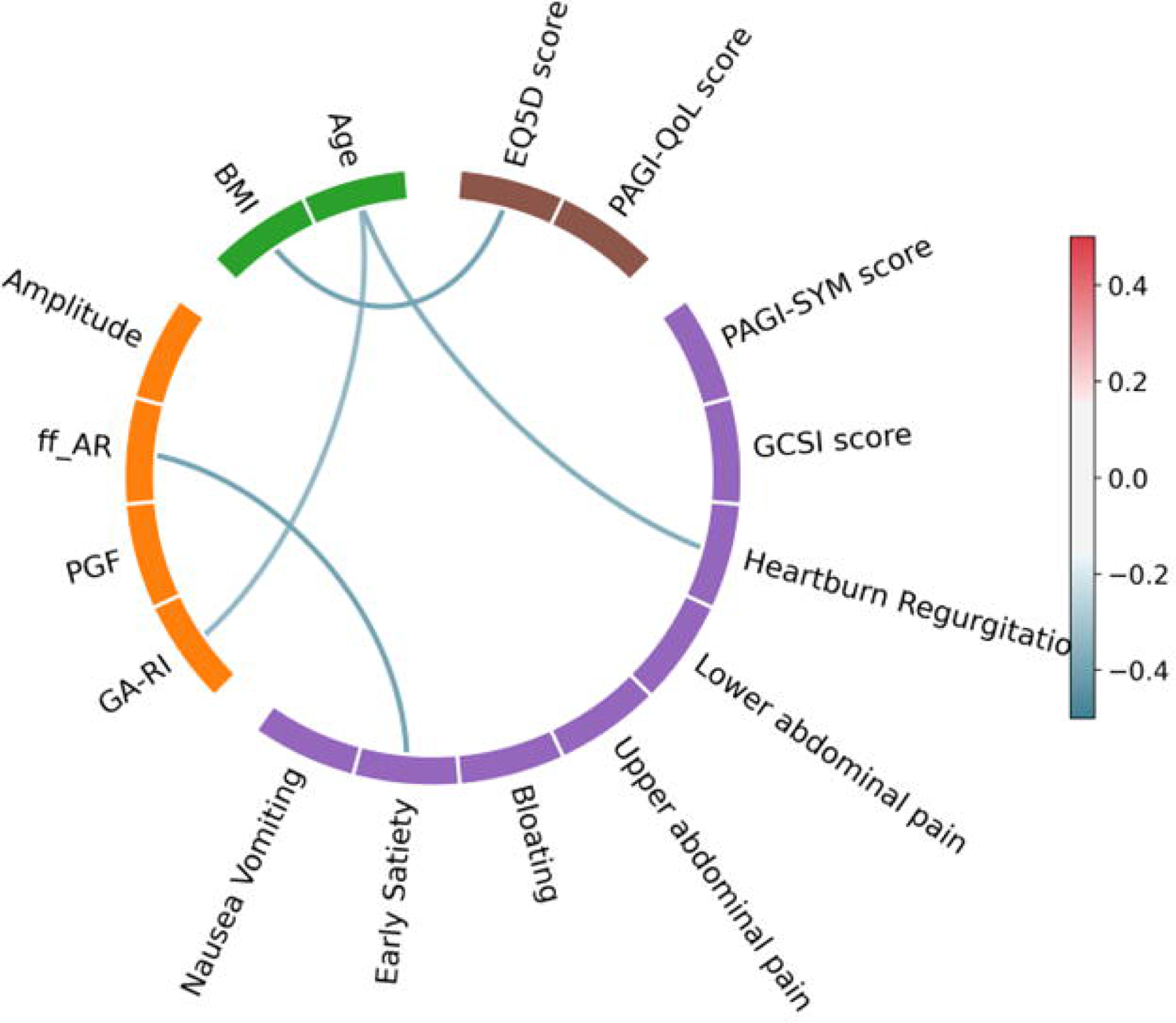

### Safety Outcomes

No adverse events were identified in any participant.

## Discussion

Chronic gastric symptoms, gastric dysfunction, and delayed gastric emptying (DGE) are recognised as common post-operative complications following pancreaticoduodenectomy (PD). While most patients improve spontaneously, a proportion experience ongoing symptoms of nausea, vomiting, reflux, thereby impacting on QoL (1, 4). This study has confirmed the safety and feasibility of the novel Gastric Alimetry system to assess the gastric function following PD. Detailed electrophysiological studies with new spectral metrics did not show significant systematic changes after PD. Post-PD patients generally also showed only modestly higher upper GI symptom burdens than controls, although QOL was substantially impaired. Gastric Alimetry symptom phenotypes were variable, demonstrating likely contributions from gastric sensorimotor, gut-brain-axis and small bowel causes all present in the cohort, which may contribute to symptom genesis.

Surgery on the stomach has been known to affect the native electrophysiology and thereby affect gastric contraction and motility (6, 9, 20, 21). Despite extensive research into surgical techniques to improve the rates of DGE (eg. antecolic vs retrocolic reconstruction, pylorus-preserving resection vs pylorus-resecting resection, Roux-en-Y vs gastrojejunal reconstruction), the evidence is still conflicting and there has been very little robust research into the underlying electrophysiology of the remnant stomach (2, 22). Recent advances in gastric mapping, including the high resolution intra-operative serosal mapping technique and the high-resolution non-invasive Gastric Alimetry system has led to significant improvements in the ability to quantify the electrophysiological changes of the stomach following surgery. Procedures such as sleeve gastrectomy, Billroth I reconstruction, and esophagectomies have all now been studied, with abnormal electrophysiology being identified in these cohorts (7, 10, 23). The Gastric Alimetry system not only employs a high-resolution array to accurately measure the gastric electrical activity, but also concurrently records the patients’ symptom evolution and defines associated objective phenotypes that may localise symptom origins within the GI tract, thereby enabling accurate correlation between symptoms and abnormal electrophysiology. This study is the first to evaluate post-PD gastric physiology and symptom profiles using the new Gastric Alimetry system.

The Gastric Alimetry spectral results showed minimal electrophysiological changes after PD, despite the terminal antral resection in the high majority of included patients, indicating that successful remodelling of gastric function occurs in most cases. While 8/19 patients had at least 1 abnormal spectral characteristic, these were generally minor, lying just outside the reference intervals. This included BMI-adjusted amplitude, which remained similar to the control group, which is of technical interest because distal antral activity has sometimes been assumed to contribute most to the body surface electrical signal (24, 25), due to the closer proximity of the antrum to the epigastrium and the high-amplitude waveforms of the terminal antral contraction (26). However, the large Gastric Alimetry array likely compensates for this by also comprehensively measuring electrical waves propagating throughout the corpus and upper antrum (14, 27). Interestingly, three cases showed a high BMI-adjusted amplitude, which could potentially be related to increased gastric contractility or an altered approximation of the stomach to the epigastric skin postoperatively due to adhesions.

There were also 2 cases exhibiting an elevated Principal Gastric Frequency. The cause of this is unclear, but this could conceivably be due to either random statistical chance given that the abnormalities were mild, or abnormal conduction of the higher frequency intestinal waves propagating retrograde into the gastric remnant, leading to the ‘Gastrointestinal Aberrant Pathway’ (GAP Syndrome) (6). This interesting possibility could be resolved in future by introducing further spatial analytics in future, which are capable of assessing the direction of gastric electrical wave propagation in the human stomach (28, 29). In the GAP abnormality, waves would be expected to travel retrograde in association with elevated frequencies (6), and in the isolated reports to date, symptoms appear to be more severe than those present in the current cohort.

When compared to the matched cohort, there were no significant differences in the spectral values observed. Only three patients were found to have concurrent spectral abnormalities and moderate to severe symptoms during this test with no consistency in the type of spectral abnormality. This finding, together with the mild nature of the electrophysiological disturbances, indicates that other factors are likely responsible for these patients’ chronic symptoms.

The range of symptom phenotypes observed in this study demonstrated the emerging capabilities of the Gastric Alimetry system to localize the source of symptom origins within the GI tract (14). Symptom origins were found to arise from a range of sources, including gastric sensorimotor symptoms (likely reflecting either reduced accommodation and/or hypersensitivity (30), post-gastric symptoms (possibly from small bowel distension or dumping (14), or continuous symptoms that could relate to post-operative pain syndromes or neuropathy (14, 19). Symptom localisation in this manner could contribute to directing therapies to improve quality of life in affected patients in future.

While the Gastric Alimetry spectral analyses did not resolve symptom causation in this cohort studied in the years after PD, gastric emptying status was not assessed. Further research is therefore now needed to assess whether changes in gastric electrophysiology might occur earlier in the post-operative course, as previously found with legacy electrogastrography techniques (3, 6), which could plausibly resolve a contributing factor to the specific complication of DGE (31).

It is well known that patient QoL is poorer following PD, which can be multifactorial, including due to chemotherapy or poor prognosis (4, 20). Of particular note, our patient cohort was found to have a significantly worse PAGI-QOL, which specifically resolves QOL related to upper GI symptoms, and self-perceived EQ-5D-5L scores compared to the matched cohort (p<0.001). No correlations, however, were identified between spectral characteristics and patient’s individual symptoms and QOL scores.

The strength of this study includes the use of a state-of-the-art non-invasive technique to assess the gastric electrophysiology of the stomach, together with standardized app and questionnaire based symptom and QoL assessments. The main limitation of the study is the number of patients recruited to identify an effect. This is particularly relevant to the occurrence of ‘GAP Syndrome’ (aberrant conduction pathway from small intestine to stomach), which has been robustly defined after Billroth I reconstruction using invasive mapping (7), and therefore appears to be a relatively rare complication. Importantly, this study also only assessed long-term changes after PD. Future studies in assessing gastric electrophysiology and motility pre- and post-surgery will also enable research into the effect of different operative variations and also potentially allow surgeons to predict patients at risk of developing DGE in the early post-operative course. Due to the complexity of the gastric reconstruction following PD, modifications to the normal Gastric Alimetry protocol were completed. This included a CT-guided placement of the array over the stomach and a reduction in size of the standard ‘meal challenge’, to 50%, accounting for the reduced gastric volume. Despite the decreased meal size, the caloric content is still considered to be adequate in stimulating gastric activity (7). Due to these modifications, patient data were compared to both the validated reference range and also a matched healthy control cohort, who also received half the caloric content compared to the standard ‘meal challenge’ (15, 16, 32).

In conclusion, this study is the first to assess the gastric electrical activity of the remnant stomach following pancreaticoduodenectomy using the new Gastric Alimetry System. Abnormal gastric electrical activity was identified, but changes were typically mild and did not correlate with symptoms or QOL, indicating that other factors contribute more to long-term QOL outcomes after PD.

## Data Availability

All data produced in the present study are available upon reasonable request to the authors

## Acknowledgements

We thank the volunteers who participated in this research and our Auckland clinical research coordinators Gen Johnston and India Wallace.

